# Association between maternal depression and the nutritional status of children under five-years-old in Peru: An analysis of the Demographic and Health Survey 2014-2017

**DOI:** 10.1101/2021.10.31.21265721

**Authors:** Nicole Villagaray-Pacheco, Pamela Villacorta-Landeo, Leslie Mejía-Guerrero, Manuel A. Virú-Loza, Percy Soto-Becerra

## Abstract

**Objective:** To evaluate the association between maternal depression and nutritional status of children under 5 years old in Peru.

**Design:** Cross-sectional study of the Demographic and Health Survey 2014-2017. Outcomes were mild (height Z-score for age <-2 SD) and moderate/severe (<-3DE) childhood chronic undernutrition and also mild (hemoglobin <11 g/dL) and moderate/severe (hemoglobin <10 g/dL) childhood anemia. Maternal depression was assessed by the questionnaire PHQ-9. Odds ratios and their confidence intervals (CIs) were estimated by multinomial logistic regression models, considering the complex sample design.

**Setting:** Peru

**Participants:** Peruvian women of childbearing age from 15 to 49 years who live with children from 6 to 59 months.

**Results:** Maternal depression was significantly associated with a higher odds of moderate/severe chronic undernutrition in children aged 6-59 months (OR = 2.67; 95% CI 1.16-6.16).

**Conclusions:** There was evidence that maternal depression was associated with an increased risk of moderate/severe chronic undernutrition

## Introduction

Malnutrition originates from an imbalance in the nutritional status of people, either due to a deficiency or excesses in caloric or nutrient intake. This includes anemia and undernutrition mainly^(1)^. Anemia and childhood undernutrition are public health problems of global importance, which prevent optimal cognitive and social development^(2–4)^. In Peru, the prevalence of child undernutrition was 13.1% in 2016, and that of childhood anemia was 43.6% in 2016 and 2017^(5,6)^.

Interventions to reduce the prevalence of childhood anemia and undernutrition have a great economic impact in Peru. The budget to combat anemia and undernutrition is between 2 165 to 3 105.99 million soles^(7,8)^. The interventions achieved a 58% reduction in infant mortality associated with undernutrition^(9)^. However, some studies suggest that interventions should be complemented with the evaluation and improvement of psychosocial determinants of maternal health, empowerment and psychological state^(10,11)^.

Maternal mental health is a determining factor in the nutritional and neurological development of children during the first years of life^(12)^. Studies reveal that children’s nutritional status is influenced by maternal mental health^(13–16)^, since its impairment decreases the ability to provide adequate care to their children and develops negative parenting behaviors^(17,18)^.

Maternal depression in developing countries affects between 19% and 25% of mothers^(19)^. In Peru, it is estimated that perinatal depression represents between 24% and 40% but only 10% of this population receives care^(19)^. As maternal depression is treatable, it could be part of population-level interventions^(19,20)^. In addition, it is an important objective in programs that seek to reduce childhood anemia and undernutrition^(18–21)^.

Several studies have found an association between maternal mental health and childhood nutritional status^(13–16)^. However, not all studies arrive at the same conclusions^(22,23)^. For this reason, the aim of our study is to determine the association between maternal depression and infant nutritional status in Peru. Studies of this type in Peru are necessary to evaluate the possibility of country-level interventions.

## Methods

### Design and study population

The study used cross-sectional data from the Demographic and Health Survey (DHS) in Peru between 2014 and 2017, using a probabilistic, stratified, independent and two-staged sample. In the first stage conglomerates were selected, while in the second, dwelling selection was performed. In both urban and rural areas, conglomerates were distributed. Conglomerates were geographic areas made up of one or several blocks that include an average of 140 households each, with a selection probability proportional to their size. Between 2014 and 2017, the annual number of households ranged from 29,941 to 35,919; while the unweighted sample sizes of women aged 15 to 49 years were 16 000 per year on average, and the weighted sample sizes of children aged 6 to 59 months were 3 000 per year on average^(24)^.

#### Maternal Depressive symptoms

The exposure of interest was maternal depressive symptoms in the last two weeks determined by a Patient Health Questionnaire-9 (PHQ9) score ≥ 10 points^(25)^. This questionnaire has nine items with scores from 0 to 3. A 10 cut-off point has a sensitivity of 88% and a specificity of 92%^(26)^.

#### Childhood chronic undernutrition

Chronic undernutrition in children was defined according to the World Health Organization (WHO) criteria as mild (Height-for-age z-score < -2 standard deviations) and moderate/severe (Height-for-age z-score < -3 standard deviations).

#### Childhood anemia

Childhood anemia was determined by the colorimetric method with a HemoCue7#x25A1; portable equipment (HemoCue AB, Angelhome, Sweden), which has a precision similar to direct methods with venous and arterial blood^(27)^. According to the WHO criteria, it was classified as mild (Hemoglobin <11g/dL) and moderate/severe (Hemoglobin <10g/dL).

#### Covariates

Confounding variables related to the child were: sex, age, birth weight, disability and health insurance; and those related to the mother were: age, pregnancy, number of pregnancies, body mass index (BMI), anemia, health insurance, marital status, ethnicity, education level, work and home status, natural region, area of origin, economic level and access to the “Programa Juntos” (a government’s economic support program). Another variable was the year in which the survey was carried out.

#### Ethical approval

The data were obtained from the open-access website of the National Institute of Statistics which is responsible for the collection of the data with the previous approval of its Ethics Board.

### Analysis of data

Data analysis was performed using Stata v.14.0 statistical package. Data-sets from the 4 survey rounds were pooled. Sample weights were used to account for uneven sampling probability and non-response. The descriptive analysis of the confounding variables was performed using absolute frequencies and percentages. Bivariate analysis for categorical variables was performed using the Chi Square test corrected for complex sample design. Numerical variables were compared between groups using the Wald test corrected for complex sampling. Multilevel analyses represented the strata and sample groupings. Logistic regression analyses included potential confounding variables, and the crude (cORm) and adjusted (aORm) multinomial Odds Ratios were estimated with a 95% confidence interval (CI) and considering the complex sample design.

## Results

### Population characteristics

A total of 11,518 children with an average age of 32.6 ± 15.7 months were included. The time period was between 2014 to 2017. Regarding sex, 51.7% were males and 48.2% were females. Regarding birth weight, 7.2% had low birth weight (<2500g). The average age of the 11,518 mothers was 26.1 ± 5.6 years. Regarding location by geographic region, they were distributed in Metropolitan Lima and the rest of the Coast (55.6%), Highlands (29.6%) and the Jungle (14.8%). 76% and 24% lived in urban and rural areas, respectively (Table 1).

**Table 1.**
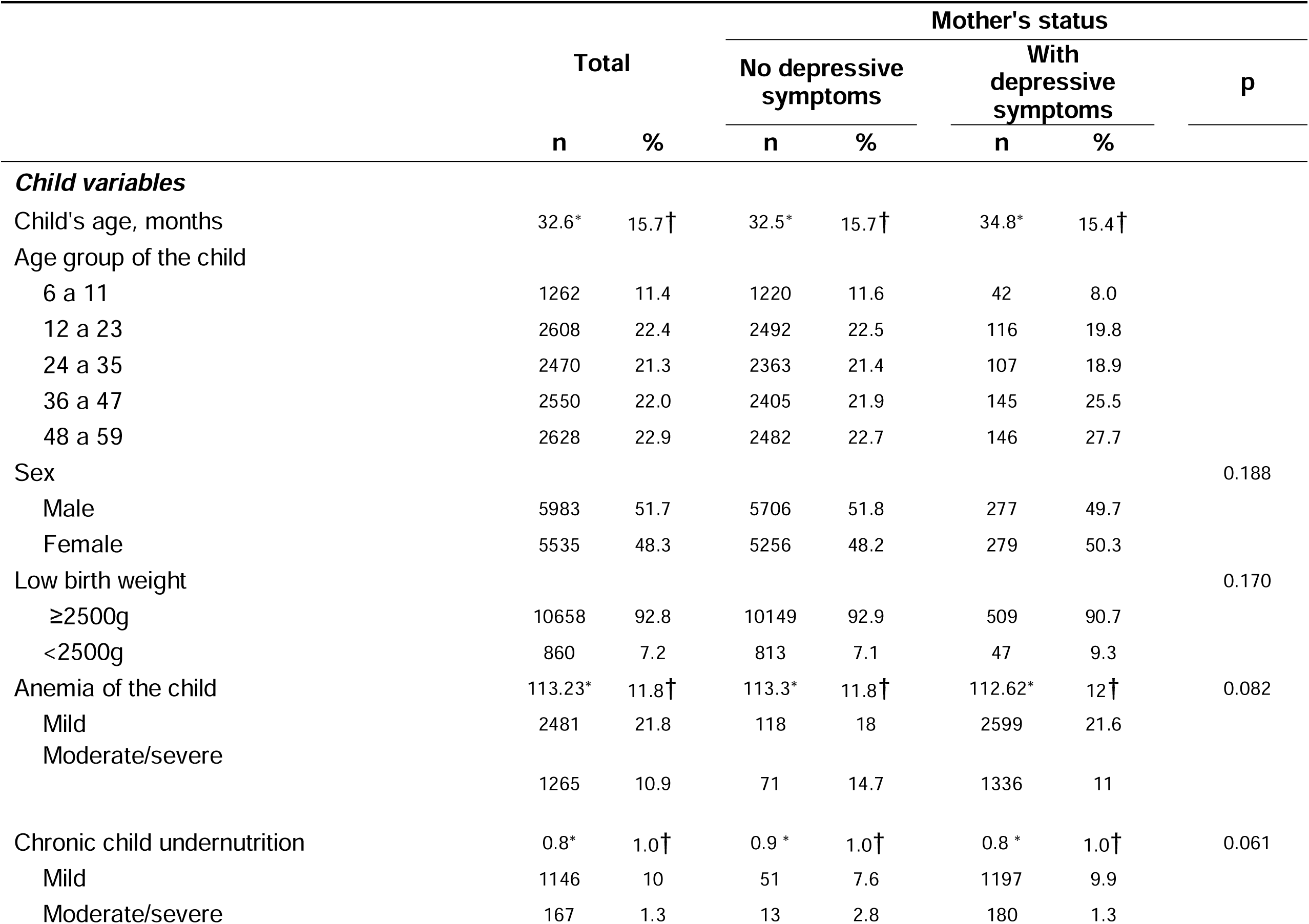
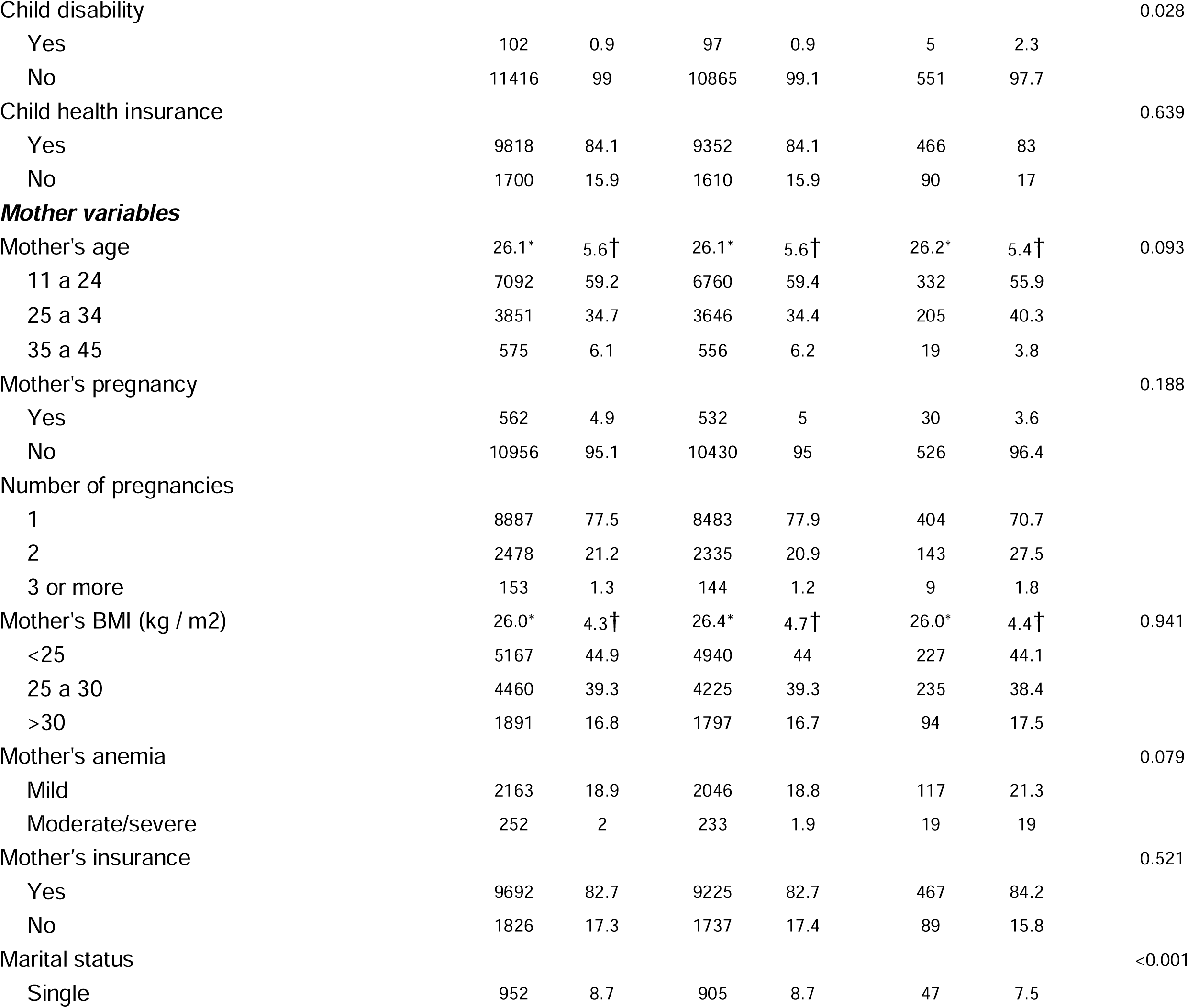
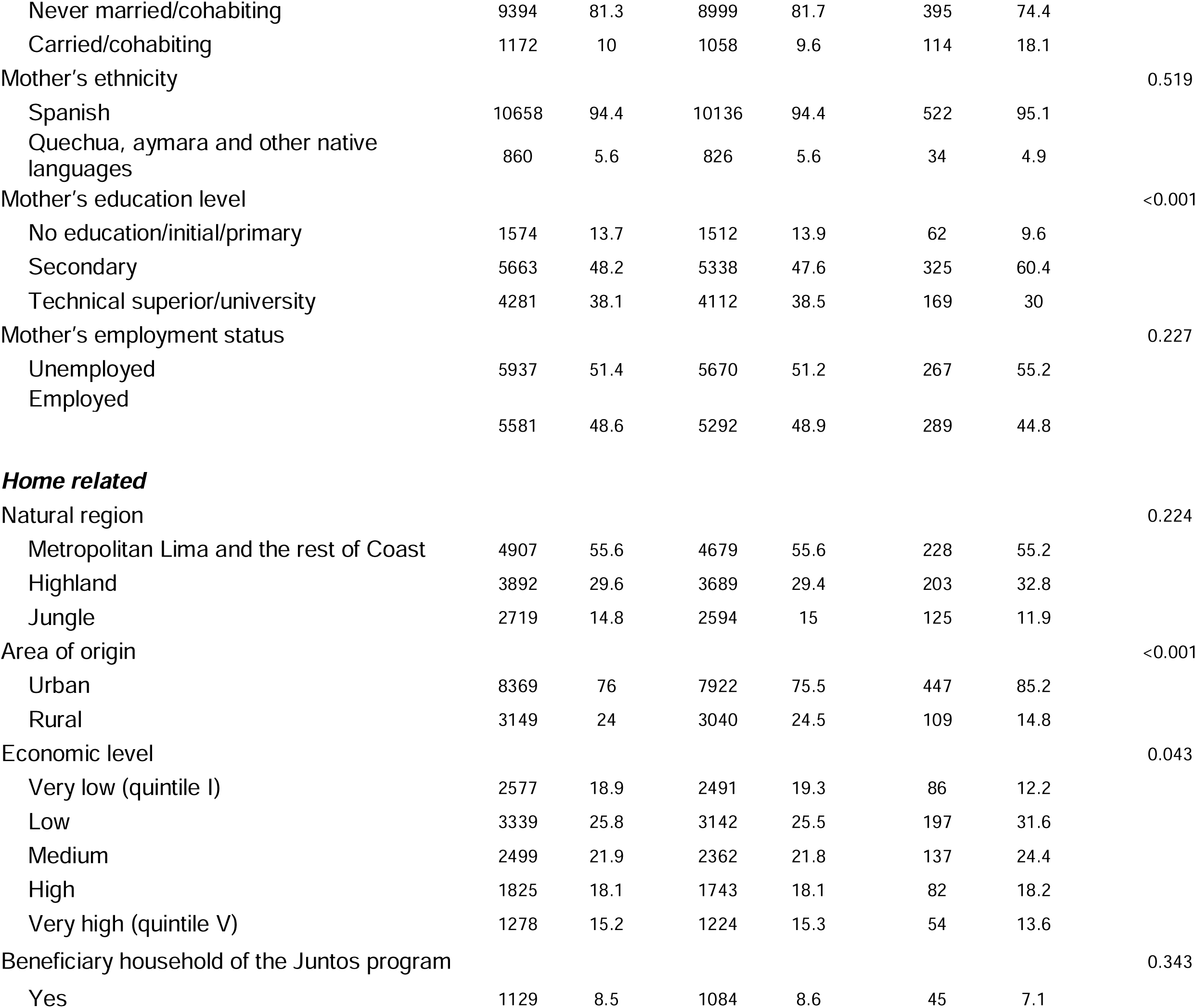
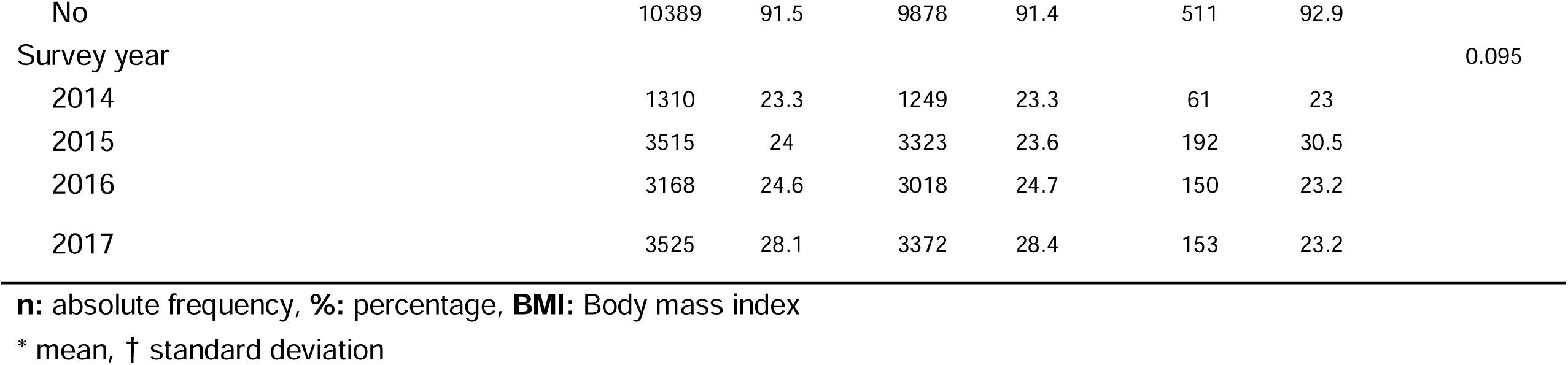
Characteristics of children under five years-old according to depressive symptoms in their mothers (Demographic and Health Survey 2014-2017)

### Prevalence of anemia and chronic undernutrition in children aged 6-59 months

The prevalence of anemia in children by age was 57%, 50%, 29.2%, 21.5% and 18% in those aged 6-11 months, 12-23 months, 24-35 months, 36-47 months and 48-59 months, respectively. Regarding the prevalence of anemia by sex (p-value=0.033), we found in males that 22% had mild anemia and 11.9% moderate/severe anemia. On the other hand, in females, 21.2% and 10.1% had mild and moderate/severe anemia, respectively (Table 2).

**Table 2.**
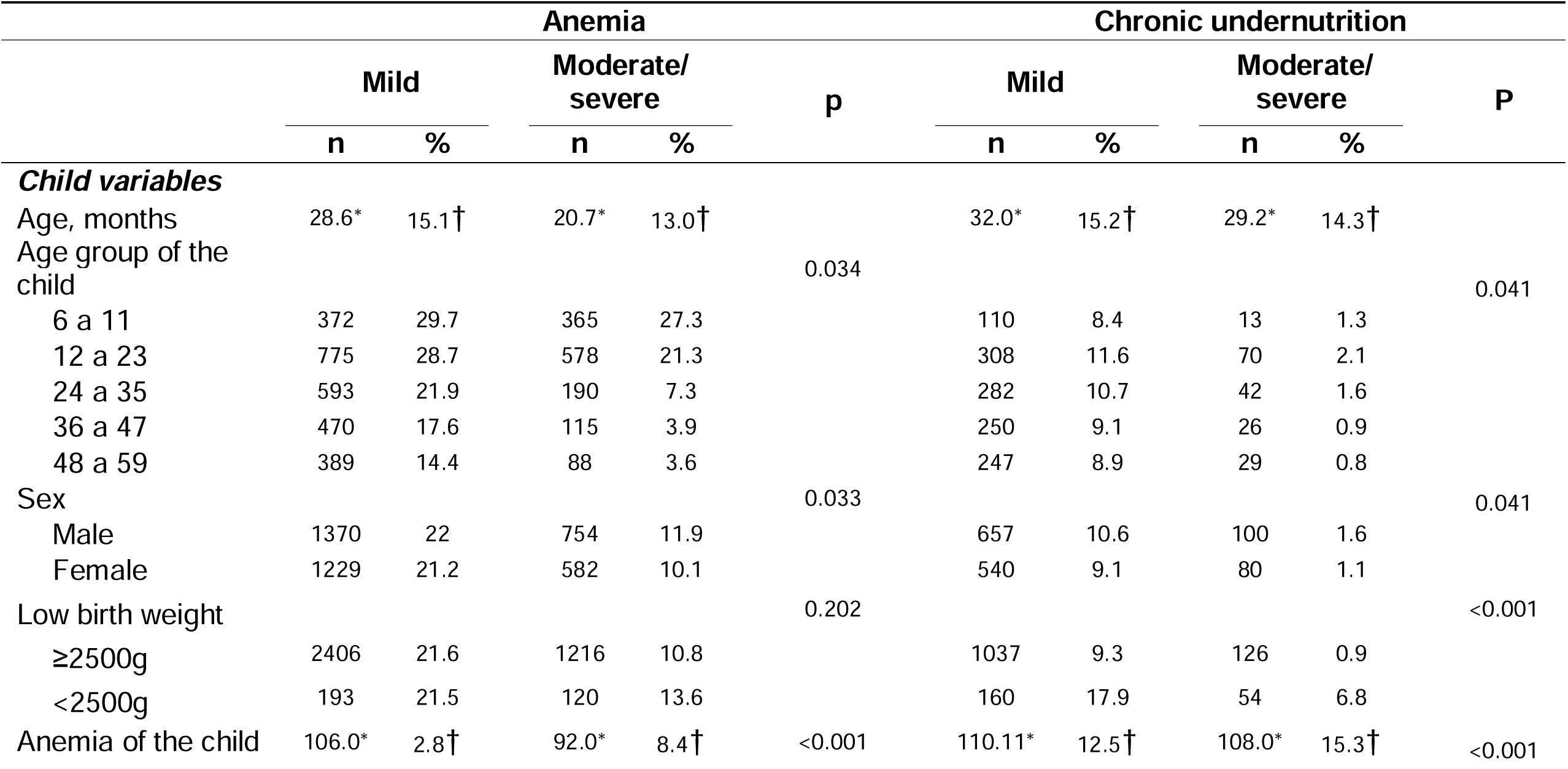
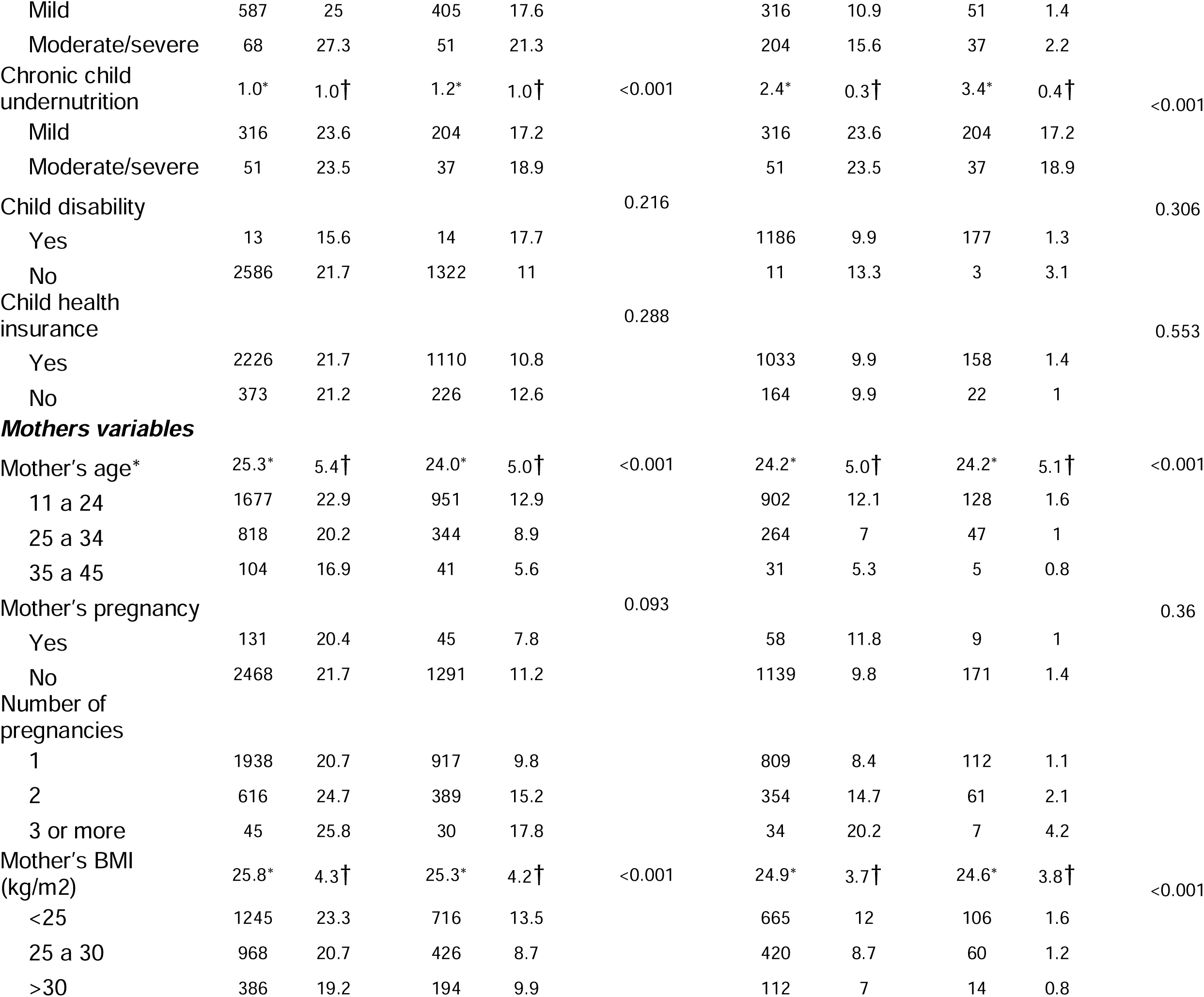
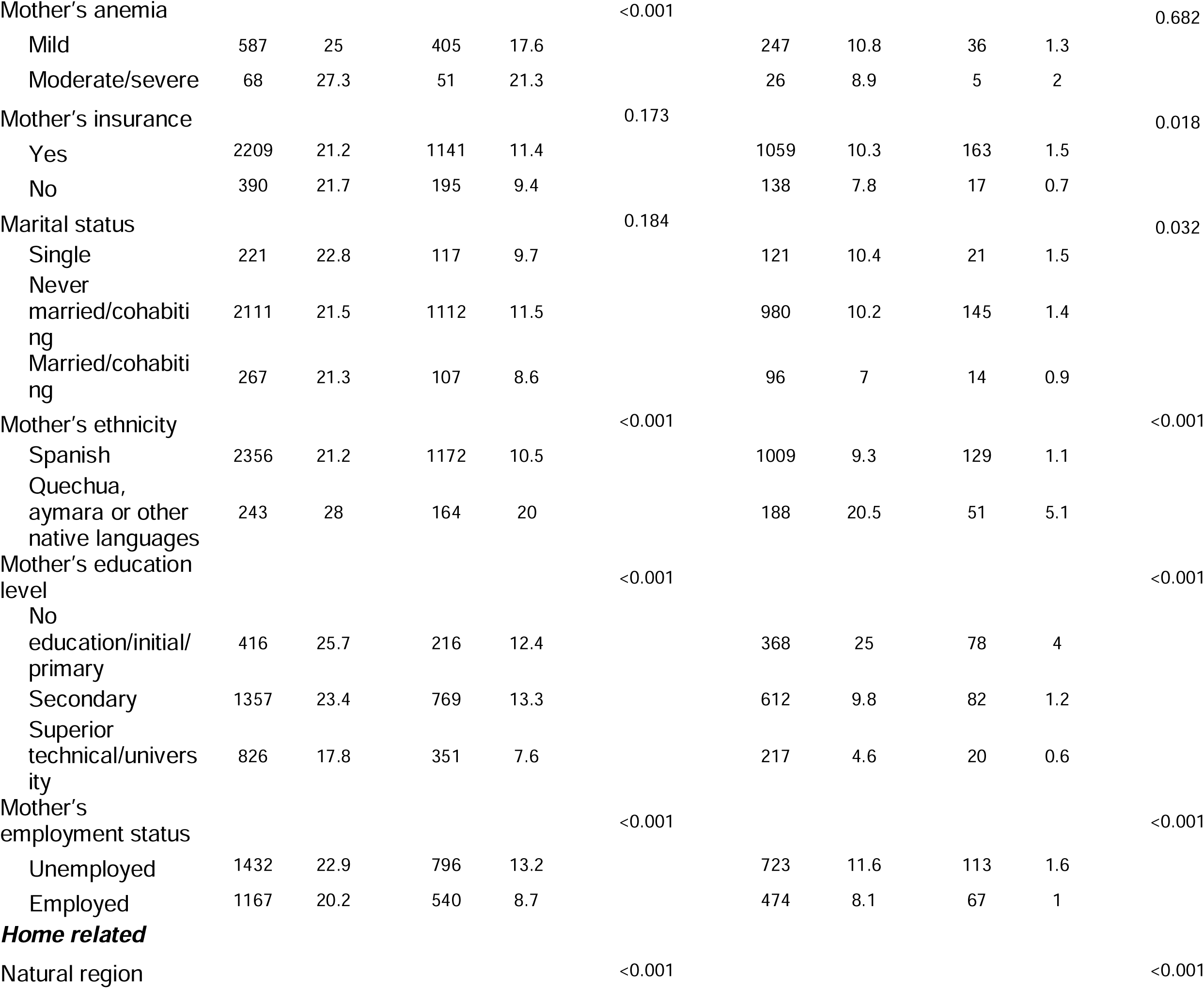
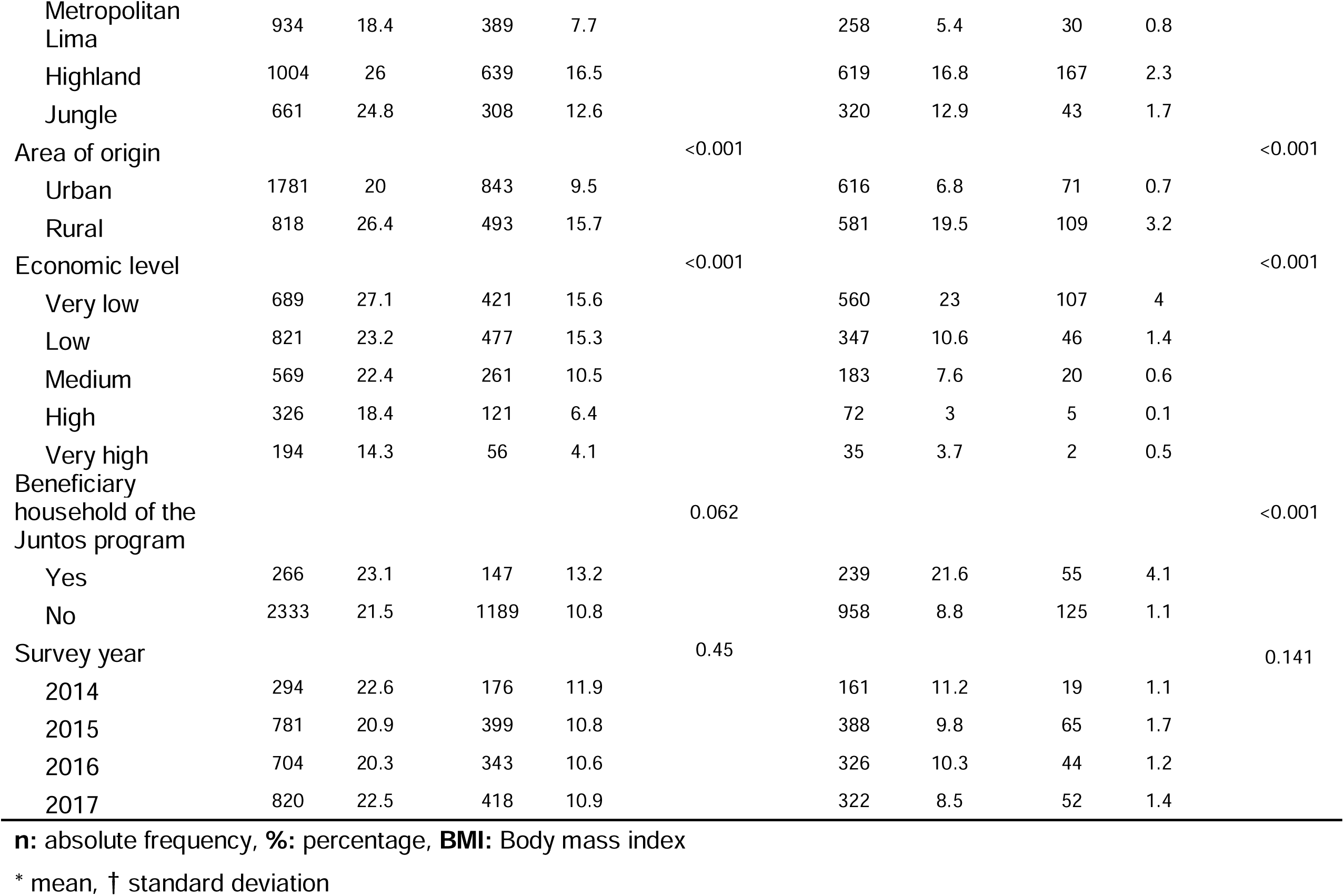
Prevalence of anemia and chronic undernutrition in children under five years-old and its bivariate association with selected characteristics (Demographic and Health Survey 2014-2017)

The prevalence of chronic undernutrition in children by age was 9.7%, 13.7%, 12.3%, 10% and 9.7% in those aged 6-11 months, 12-23 months, 24-35 months, 36-47 months and 48-59 months, respectively. Regarding the prevalence of chronic undernutrition by sex (p-value=0.041) we found in males that 10.6% had mild chronic undernutrition and 1.6% had moderate/severe chronic undernutrition. On the other hand, in females, 9.1% and 1.1% had mild and moderate/severe chronic undernutrition, respectively (Table 2).

### Mother’s depressive symptoms and anemia in children aged 6-59 months

In children, neither mild anemia (ORm=0.082; 95% CI; p = 0.264) nor moderate/severe anemia (ORm=1.36; 95% CI; p = 0.086) were significantly associated with mother’s depressive symptoms in crude analysis. Adjusted analysis led to the same conclusion for mild (ORm = 0.87; 95% CI; p = 0.445) and moderate/severe anemia (ORm = 1.53; 95% CI; p = 0.058) (Table 3).

**Table 3.**
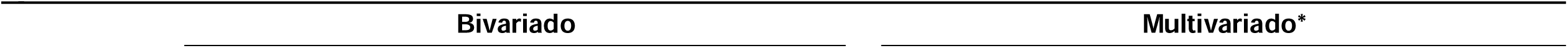
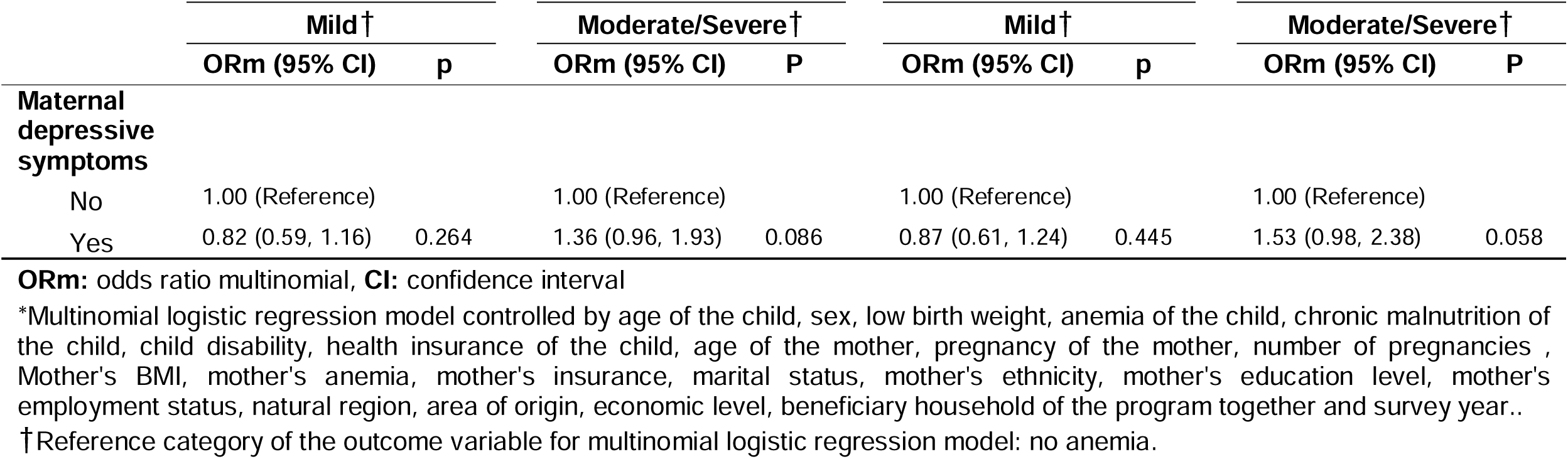
Adjusted and unadjusted association between maternal depressive symptoms and anemia in children under five.

### Mother’s depressive symptoms and chronic undernutrition in children aged 6-59 months

In children, neither mild chronic undernutrition (ORm = 0.76; 95% CI; p = 0.177), nor moderate/severe chronic undernutrition (ORm = 2.19; 95% CI; p = 0.080) were significantly associated with mother’s depressive symptoms in unadjusted analysis. Adjusted analysis led to the same conclusion for mild chronic undernutrition in children (ORm = 0.81; 95% CI; p = 0.372). However, moderate/severe chronic undernutrition showed a significant association (ORm = 2.67; 95% CI; p = 0.0021) (Table 4).

**Table 4.**
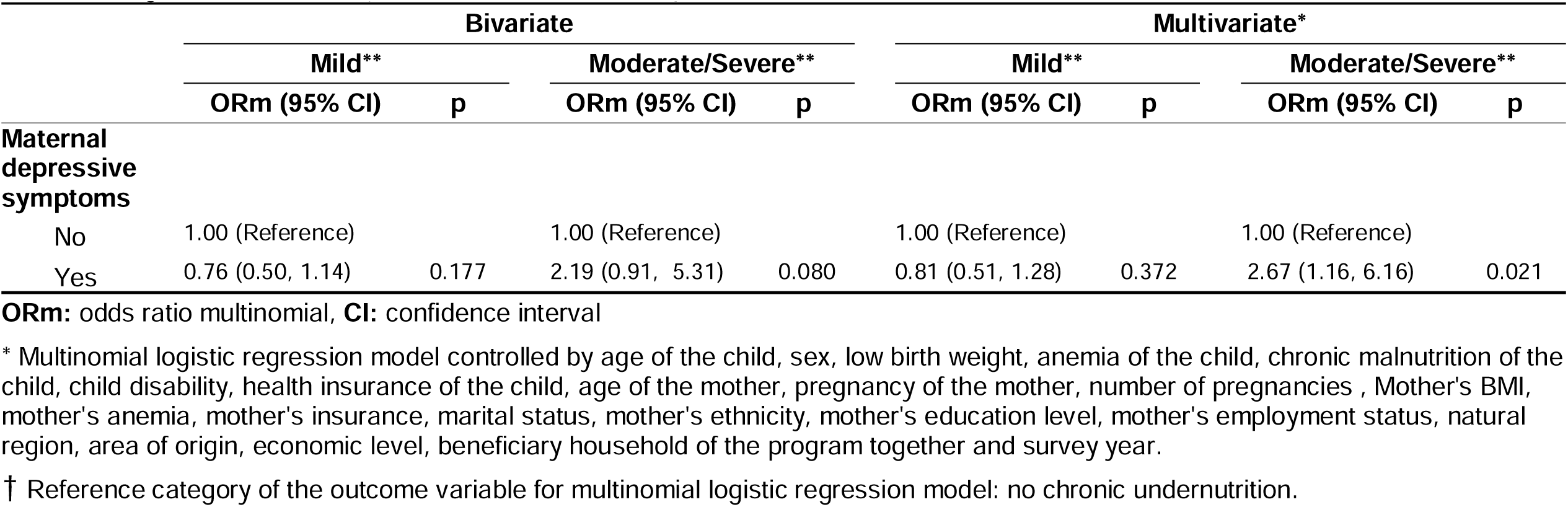
Adjusted and unadjusted association between maternal depressive symptoms and chronic undernutrition in children under five years-old (Demographic and Health Survey 2014-2017)

## Discussion

During the 4-year period evaluated, the present study found that children of mothers who have depression have 2.67 times more probability to have moderate/severe chronic undernutrition than those who do not have mothers with depression. This is the first publication on the association between maternal depression and the nutritional status of children in a period of 4 years with a representative sample. Our results suggest that the late diagnosis and treatment of maternal depression is related with a moderate/severe poor nutritional status of children.

The highest national prevalence of chronic undernutrition in children between 6 to 59 months was 14.6% in 2014, but in later years this figure was decreasing^(28)^. Compared to WHO global figures, national figures are 2 times higher^(28,29)^. There was a higher prevalence in those living in urban areas, belonging to the highlands and being in the bottom quintile of wealth, according to an analysis by subgroups^(30)^. The highest national prevalence of anemia in children aged 6 to 35 months between 2014 and 2017 was 46.8% in 2014. This was 2 times higher compared to WHO global figures^(28–31)^. According to subgroup analysis, moderate and severe anemia were the most prevalent^(32)^.

Regarding undernutrition, Ashaba et al. carried out a study in children between 1 and 5 years of age in Africa, in which they concluded a 2.4 times greater probability of undernutrition in children of depressed mothers compared to children of mothers without depression (OR: 2.4, 95% CI: 1.1 to 5.18, p: 0.03), but this study had a short follow-up and few measurements^(33)^. Anoop et al. carried out a study in children of mothers with symptoms of postpartum major depression in India, in which children were evaluated in the 6th, 10th, 14th, 36th and 72nd weeks after birth, that suggests a 7.4 times greater probability of presenting child undernutrition compared to children of mothers without symptoms (OR: 7.4, 95% CI: 1.6 to 38.5, p: 0.01)^(34)^. The association was 3 times that found in our study. This may be due to its longitudinal design, so it had multiple measurements to monitor the child’s nutritional status. In addition, it was carried out in a rural area, where there is a greater probability of presenting depression due to the difficulty of accessing health services^(35)^. Mohatlhedi et al. conducted a case-control study in Botswana, where they assessed the association between depression in non-maternal primary caregivers and malnourishment in children from 6 months to 5 years of age^(36)^. They found a 4.3 times greater presence of malnutrition in children with depressed caregivers compared to children with non-depressed caregivers (OR = 4.33, 95% CI: 1.90 to 9.90, p: 0.001). The association was 2 times that found in our study. This could be due to the sociodemographic characteristics of the caregivers since a lower educational level and wealth are associated with less care provided to children^(36)^. Furthermore, the care provided by caregivers may be less than that provided by mothers.

Regarding chronic undernutrition, Nguyen et al. published a study with children between 19 and 42 months of age and their respective mothers in India, in which the authors suggest a 1.4 times greater probability of presenting growth retardation in children with mothers with high depression indexes (OR: 1.47, 95% CI: 1.09 to 1.98, p <0.05)^(37)^. This association is less than that in our study and may be due to the fact that most of those mothers were housewives and spent more time caring for their children, as well as belonging to ethnic groups where there is an important role for the mother in taking care of their children. Surkan et al. carried out a study in 3 Brazilian cities, whose population consisted of 595 low-income mothers and their respective children aged 6 to 24 months^(38)^. They concluded that maternal depressive symptoms increased the chances of delaying child growth in 1.8 times (OR: 1.8, 95% CI: 1.1 to 1.98, p <0.01). Although the sample size is smaller than ours, the results are significant and can be generalized. Rahman et al. conducted a prospective cohort study in Pakistan, which included 160 depressed mothers, 160 psychologically healthy mothers, and their respective children^(39)^. The results suggested that maternal depression increased the chances of stunting by 4.4 times in 6-month-old children (RR: 4.4, 95% CI: 1.7 to 4.1) and 2.5 times in 12-month-old children (RR: 2.5, 95% CI: 1.6 to 4.0). The association in 12-month-old children is similar to that found in our study since their study sample had characteristics similar to ours such as the age of mothers, which is between 17 to 40 years old; however, the study designs are different.

On the other hand, some studies did not demonstrate association between maternal depression and child undernutrition, when it was analyzed by sociocultural variables. For example, Joshi, et al. published a study with 300 children between 0 to 11 months and their mothers in India. They concluded that maternal depression did not present a significant association with nutritional status indicators like stunting (OR: 1.485, 95% CI: 0.833 – 2.649)^(40)^. The results differ from ours, maybe because the family characteristics are different since the majority of mothers surveyed had a nuclear and complete family, so they probably received emotional and financial support from their husbands. Emerson et al. carried out a study with 828 children and their mothers, who participated in Congo’s polls. Their study concluded that being underweight in children (OR: 0.91, 95% CI: 0.60 to 1.37, p=0.64) did not have a significant association with maternal depression^(41)^. The results are different from ours despite similar socio-cultural characteristics study populations. This could be because their study population lived in a place with diverse food resources and provided high levels of nutrients.

There is no precedent of studies that had evaluated directly the association between maternal depression and anemia; therefore, we will discuss studies that determined the association between anemia and inadequate mother-child interaction. For example, Corapci et al. conducted a 5-year longitudinal study in Costa Rica, which included babies between 12 and 23 months of age and their respective mothers, that suggested a significant association between a lower quality of mother-child interaction with chronic iron deficiency (OR: 0.43, 95% CI: 0.22 to 0.87, p <0.05)^(42)^. These results differ from the marginally significant association that we found, which may be because some maternal socio demographics characteristics are different such as the maternal economic and education level. In the study mentioned mothers have a very low-low economic level and only reached incomplete high school education^(42)^. However, in our study, the maternal economic and education level are mainly low-medium and high-school or higher education, respectively.

According to Pullum et al. DHS in Peru faces methodological challenges^(43)^. The data provided by the mothers are self-reports of their health, their own experience in caring for their children. Only anthropometric measurements are obtained independently and are not affected by self-report bias. Regarding anthropometry, important sources of error include incorrect measurement of age and height/length. DHS reports the digit bias in the child’s height record but concludes that the digit bias will probably not introduce a significant level of error in the calculation of the chronic undernutrition variables. However, to reduce this type of error, a series of procedures were carried out, such as careful design and numerous tests of the questionnaire, intense training of the interviewers, arduous supervision and permanent fieldwork, revision of the questionnaires, appropriate supervision at the coding and processing of data and careful cleaning of the file with feedback to supervisors and interviewers from quality^(44–46)^.

The operational definition of maternal depression deserves critical consideration. The depressive symptoms are not accurate enough to diagnose depression. However, it can be measured in an indirect way using depressive symptoms. The standard used for depressive symptoms is the PHQ-9 survey, which was validated in Peru^(25)^, since it has many advantages as it is a rapid screening instrument, self-applied, and a near diagnostic tool. Furthermore, it indicates the severity, so it would serve to monitor the management and evolution of symptoms. On the other hand, the standards for determining chronic undernutrition and anemia were established by the WHO based on the results of the Multicentre Growth Reference Study^(47,48)^. Therefore, the general pattern of results seems to reasonably support our main conclusions.

Despite these limitations, our study provides unique data on the association between maternal depression and poor child nutritional status using the currently recommended standards for chronic undernutrition, anemia and depression, and provides estimates for a large demographic group. It shows evidence that maternal depression is significantly associated with chronic undernutrition. Besides, we found a marginally significant association between maternal depression and anemia. Furthermore, the results of this study will contribute to increasing maternal mental health research, which is a research priority in Peru^(49)^. Likewise, they could be useful for the development of prevention policies against anemia and undernutrition in children of mothers with depressive syndrome by organizations such as WHO, Pan American Health Organization (PAHO), United Nations International Children’s Emergency Fund (UNICEF) and the Peruvian Ministry of Health. In addition, the results will help to promote prevention policies related to maternal mental health, because the decline of it could be harmful to the nutritional status of their children.

## Supporting information

Table 1, and will be used for the file on the preprint site

Table 2, and will be used for the file on the preprint site

Table 3, and will be used for the file on the preprint site

Table 4, and will be used for the file on the preprint site

## Data Availability

All data produced are available online at http://iinei.inei.gob.pe/microdatos/

## Notes

### Competing Interest Statement

The authors have declared no competing interest.

### Funding Statement

This study did not receive any funding

### Author Declarations

This study involves only openly available human data, which can be obtained from: http://iinei.inei.gob.pe/microdatos/

