## Supplementary material for "Association between maternal depression and the nutritional status of children under five-years-old in Peru: An analysis of the Demographic and Health Survey 2014-2017": Table 1, and will be used for the file on the preprint site

| **Table 1. Characteristics of children under five years-old according to depressive symptoms in their mothers (Demographic and Health Survey 2014-2017)** | | | | | | | | | | | | |
| --- | --- | --- | --- | --- | --- | --- | --- | --- | --- | --- | --- | --- |
|  |  | **Total** | | |  | **Mother's status** | | | | | | |
|  |  |  |  |  |  | **No depressive symptoms** | |  | **With depressive symptoms** | |  | **p** |
|  |  | **n** | **%** | |  | **n** | **%** |  | **n** | **%** |  |  |
| ***Child variables*** | |  |  | |  |  |  |  |  |  |  |  |
| Child's age, months | | 32.6* | 15.7† | |  | 32.5* | 15.7† |  | 34.8* | 15.4† |  |  |
| Age group of the child | |  |  | |  |  |  |  |  |  |  |  |
|  | 6 a 11 | 1262 | 11.4 | |  | 1220 | 11.6 |  | 42 | 8.0 |  |  |
|  | 12 a 23 | 2608 | 22.4 | |  | 2492 | 22.5 |  | 116 | 19.8 |  |  |
|  | 24 a 35 | 2470 | 21.3 | |  | 2363 | 21.4 |  | 107 | 18.9 |  |  |
|  | 36 a 47 | 2550 | 22.0 | |  | 2405 | 21.9 |  | 145 | 25.5 |  |  |
|  | 48 a 59 | 2628 | 22.9 | |  | 2482 | 22.7 |  | 146 | 27.7 |  |  |
| Sex | |  |  | |  |  |  |  |  |  |  | 0.188 |
|  | Male | 5983 | 51.7 | |  | 5706 | 51.8 |  | 277 | 49.7 |  |  |
|  | Female | 5535 | 48.3 | |  | 5256 | 48.2 |  | 279 | 50.3 |  |  |
| Low birth weight | |  |  | |  |  |  |  |  |  |  | 0.170 |
|  | ≥2500g | 10658 | 92.8 | |  | 10149 | 92.9 |  | 509 | 90.7 |  |  |
|  | <2500g | 860 | 7.2 | |  | 813 | 7.1 |  | 47 | 9.3 |  |  |
| Anemia of the child | | 113.23* | 11.8† | |  | 113.3* | 11.8† |  | 112.62* | 12† |  | 0.082 |
|  | Mild | 2481 | 21.8 | |  | 118 | 18 |  | 2599 | 21.6 |  |  |
|  | Moderate/severe | 1265 | 10.9 | |  | 71 | 14.7 |  | 1336 | 11 |  |  |
| Chronic child undernutrition | | 0.8* | 1.0† | |  | 0.9 * | 1.0† |  | 0.8 * | 1.0† |  | 0.061 |
|  | Mild | 1146 | 10 | |  | 51 | 7.6 |  | 1197 | 9.9 |  |  |
|  | Moderate/severe | 167 | 1.3 | |  | 13 | 2.8 |  | 180 | 1.3 |  |  |
| Child disability | |  |  | |  |  |  |  |  |  |  | 0.028 |
|  | Yes | 102 | 0.9 | |  | 97 | 0.9 |  | 5 | 2.3 |  |  |
|  | No | 11416 | 99 | |  | 10865 | 99.1 |  | 551 | 97.7 |  |  |
| Child health insurance | |  |  | |  |  |  |  |  |  |  | 0.639 |
|  | Yes | 9818 | 84.1 | |  | 9352 | 84.1 |  | 466 | 83 |  |  |
|  | No | 1700 | 15.9 | |  | 1610 | 15.9 |  | 90 | 17 |  |  |
| ***Mother variables*** | |  |  | |  |  |  |  |  |  |  |  |
| Mother's age | | 26.1* | 5.6† | |  | 26.1* | 5.6† |  | 26.2* | 5.4† |  | 0.093 |
|  | 11 a 24 | 7092 | 59.2 | |  | 6760 | 59.4 |  | 332 | 55.9 |  |  |
|  | 25 a 34 | 3851 | 34.7 | |  | 3646 | 34.4 |  | 205 | 40.3 |  |  |
|  | 35 a 45 | 575 | 6.1 | |  | 556 | 6.2 |  | 19 | 3.8 |  |  |
| Mother's pregnancy | |  |  | |  |  |  |  |  |  |  | 0.188 |
|  | Yes | 562 | 4.9 | |  | 532 | 5 |  | 30 | 3.6 |  |  |
|  | No | 10956 | 95.1 | |  | 10430 | 95 |  | 526 | 96.4 |  |  |
| Number of pregnancies | |  |  | |  |  |  |  |  |  |  |  |
|  | 1 | 8887 | 77.5 | |  | 8483 | 77.9 |  | 404 | 70.7 |  |  |
|  | 2 | 2478 | 21.2 | |  | 2335 | 20.9 |  | 143 | 27.5 |  |  |
|  | 3 or more | 153 | 1.3 | |  | 144 | 1.2 |  | 9 | 1.8 |  |  |
| Mother's BMI (kg / m2) | | 26.0* | | 4.3† |  | 26.4* | 4.7† |  | 26.0* | 4.4† |  | 0.941 |
|  | <25 | 5167 | 44.9 | |  | 4940 | 44 |  | 227 | 44.1 |  |  |
|  | 25 a 30 | 4460 | 39.3 | |  | 4225 | 39.3 |  | 235 | 38.4 |  |  |
|  | >30 | 1891 | 16.8 | |  | 1797 | 16.7 |  | 94 | 17.5 |  |  |
| Mother's anemia | |  |  | |  |  |  |  |  |  |  | 0.079 |
|  | Mild | 2163 | 18.9 | |  | 2046 | 18.8 |  | 117 | 21.3 |  |  |
|  | Moderate/severe | 252 | 2 | |  | 233 | 1.9 |  | 19 | 19 |  |  |
| Mother’s insurance | |  |  | |  |  |  |  |  |  |  | 0.521 |
|  | Yes | 9692 | 82.7 | |  | 9225 | 82.7 |  | 467 | 84.2 |  |  |
|  | No | 1826 | 17.3 | |  | 1737 | 17.4 |  | 89 | 15.8 |  |  |
| Marital status | |  |  | |  |  |  |  |  |  |  | <0.001 |
|  | Single | 952 | 8.7 | |  | 905 | 8.7 |  | 47 | 7.5 |  |  |
|  | Never married/cohabiting | 9394 | 81.3 | |  | 8999 | 81.7 |  | 395 | 74.4 |  |  |
|  | Carried/cohabiting | 1172 | 10 | |  | 1058 | 9.6 |  | 114 | 18.1 |  |  |
| Mother’s ethnicity | |  |  | |  |  |  |  |  |  |  | 0.519 |
|  | Spanish | 10658 | 94.4 | |  | 10136 | 94.4 |  | 522 | 95.1 |  |  |
|  | Quechua, aymara and other native languages | 860 | 5.6 | |  | 826 | 5.6 |  | 34 | 4.9 |  |  |
| Mother’s education level | |  |  | |  |  |  |  |  |  |  | <0.001 |
|  | No education/initial/primary | 1574 | 13.7 | |  | 1512 | 13.9 |  | 62 | 9.6 |  |  |
|  | Secondary | 5663 | 48.2 | |  | 5338 | 47.6 |  | 325 | 60.4 |  |  |
|  | Technical superior/university | 4281 | 38.1 | |  | 4112 | 38.5 |  | 169 | 30 |  |  |
| Mother’s employment status | |  |  | |  |  |  |  |  |  |  | 0.227 |
|  | Unemployed | 5937 | 51.4 | |  | 5670 | 51.2 |  | 267 | 55.2 |  |  |
|  | Employed | 5581 | 48.6 | |  | 5292 | 48.9 |  | 289 | 44.8 |  |  |
| ***Home related*** | |  |  | |  |  |  |  |  |  |  |  |
| Natural region | |  |  | |  |  |  |  |  |  |  | 0.224 |
|  | Metropolitan Lima and the rest of Coast | 4907 | 55.6 | |  | 4679 | 55.6 |  | 228 | 55.2 |  |  |
|  | Highland | 3892 | 29.6 | |  | 3689 | 29.4 |  | 203 | 32.8 |  |  |
|  | Jungle | 2719 | 14.8 | |  | 2594 | 15 |  | 125 | 11.9 |  |  |
| Area of origin | |  |  | |  |  |  |  |  |  |  | <0.001 |
|  | Urban | 8369 | 76 | |  | 7922 | 75.5 |  | 447 | 85.2 |  |  |
|  | Rural | 3149 | 24 | |  | 3040 | 24.5 |  | 109 | 14.8 |  |  |
| Economic level | |  |  | |  |  |  |  |  |  |  | 0.043 |
|  | Very low (quintile I) | 2577 | 18.9 | |  | 2491 | 19.3 |  | 86 | 12.2 |  |  |
|  | Low | 3339 | 25.8 | |  | 3142 | 25.5 |  | 197 | 31.6 |  |  |
|  | Medium | 2499 | 21.9 | |  | 2362 | 21.8 |  | 137 | 24.4 |  |  |
|  | High | 1825 | 18.1 | |  | 1743 | 18.1 |  | 82 | 18.2 |  |  |
|  | Very high (quintile V) | 1278 | 15.2 | |  | 1224 | 15.3 |  | 54 | 13.6 |  |  |
| Beneficiary household of the Juntos program | |  |  | |  |  |  |  |  |  |  | 0.343 |
|  | Yes | 1129 | 8.5 | |  | 1084 | 8.6 |  | 45 | 7.1 |  |  |
|  | No | 10389 | 91.5 | |  | 9878 | 91.4 |  | 511 | 92.9 |  |  |
| Survey year | |  |  | |  |  |  |  |  |  |  | 0.095 |
|  | 2014 | 1310 | 23.3 | |  | 1249 | 23.3 |  | 61 | 23 |  |  |
|  | 2015 | 3515 | 24 | |  | 3323 | 23.6 |  | 192 | 30.5 |  |  |
|  | 2016 | 3168 | 24.6 | |  | 3018 | 24.7 |  | 150 | 23.2 |  |  |
|  | 2017 | 3525 | 28.1 | |  | 3372 | 28.4 |  | 153 | 23.2 |  |  |
| **n:** absolute frequency, **%:** percentage, **BMI:** Body mass index | | | | |  |  |  |  |  |  |  |  |
| * mean, † standard deviation | |  |  | |  |  |  |  |  |  |  |  |
