## Supplementary material for "Association between maternal depression and the nutritional status of children under five-years-old in Peru: An analysis of the Demographic and Health Survey 2014-2017": Table 2, and will be used for the file on the preprint site

| **Table 2. Prevalence of anemia and chronic undernutrition in children under five years-old and its bivariate association with selected characteristics (Demographic and Health Survey 2014-2017)** | | | | | | | | | | | | | | | | |
| --- | --- | --- | --- | --- | --- | --- | --- | --- | --- | --- | --- | --- | --- | --- | --- | --- |
|  |  | **Anemia** | | | | | | |  | **Chronic undernutrition** | | | | | | |
|  |  | **Mild** | |  | **Moderate/  severe** | |  | **p** |  | **Mild** | |  | **Moderate/ severe** | |  | **P** |
|  |  | **n** | **%** |  | **n** | **%** |  |  |  | **n** | **%** |  | **n** | **%** |  |  |
| ***Child variables*** | |  |  |  |  |  |  |  |  |  |  |  |  |  |  |  |
| Age, months | | 28.6* | 15.1† |  | 20.7* | 13.0† |  |  |  | 32.0* | 15.2† |  | 29.2* | 14.3† |  |  |
| Age group of the child | |  |  |  |  |  |  | 0.034 |  |  |  |  |  |  |  | 0.041 |
|  | 6 a 11 | 372 | 29.7 |  | 365 | 27.3 |  |  |  | 110 | 8.4 |  | 13 | 1.3 |  |  |
|  | 12 a 23 | 775 | 28.7 |  | 578 | 21.3 |  |  |  | 308 | 11.6 |  | 70 | 2.1 |  |  |
|  | 24 a 35 | 593 | 21.9 |  | 190 | 7.3 |  |  |  | 282 | 10.7 |  | 42 | 1.6 |  |  |
|  | 36 a 47 | 470 | 17.6 |  | 115 | 3.9 |  |  |  | 250 | 9.1 |  | 26 | 0.9 |  |  |
|  | 48 a 59 | 389 | 14.4 |  | 88 | 3.6 |  |  |  | 247 | 8.9 |  | 29 | 0.8 |  |  |
| Sex | |  |  |  |  |  |  | 0.033 |  |  |  |  |  |  |  | 0.041 |
|  | Male | 1370 | 22 |  | 754 | 11.9 |  |  |  | 657 | 10.6 |  | 100 | 1.6 |  |  |
|  | Female | 1229 | 21.2 |  | 582 | 10.1 |  |  |  | 540 | 9.1 |  | 80 | 1.1 |  |  |
| Low birth weight | |  |  |  |  |  |  | 0.202 |  |  |  |  |  |  |  | <0.001 |
|  | ≥2500g | 2406 | 21.6 |  | 1216 | 10.8 |  |  |  | 1037 | 9.3 |  | 126 | 0.9 |  |  |
|  | <2500g | 193 | 21.5 |  | 120 | 13.6 |  |  |  | 160 | 17.9 |  | 54 | 6.8 |  |  |
| Anemia of the child | | 106.0* | 2.8† |  | 92.0* | 8.4† |  | <0.001 |  | 110.11* | 12.5† |  | 108.0* | 15.3† |  | <0.001 |
|  | Mild | 587 | 25 |  | 405 | 17.6 |  |  |  | 316 | 10.9 |  | 51 | 1.4 |  |  |
|  | Moderate/severe | 68 | 27.3 |  | 51 | 21.3 |  |  |  | 204 | 15.6 |  | 37 | 2.2 |  |  |
| Chronic child undernutrition | | 1.0* | 1.0† |  | 1.2* | 1.0† |  | <0.001 |  | 2.4* | 0.3† |  | 3.4* | 0.4† |  | <0.001 |
|  | Mild | 316 | 23.6 |  | 204 | 17.2 |  |  |  | 316 | 23.6 |  | 204 | 17.2 |  |  |
|  | Moderate/severe | 51 | 23.5 |  | 37 | 18.9 |  |  |  | 51 | 23.5 |  | 37 | 18.9 |  |  |
| Child disability | |  |  |  |  |  |  | 0.216 |  |  |  |  |  |  |  | 0.306 |
|  | Yes | 13 | 15.6 |  | 14 | 17.7 |  |  |  | 1186 | 9.9 |  | 177 | 1.3 |  |  |
|  | No | 2586 | 21.7 |  | 1322 | 11 |  |  |  | 11 | 13.3 |  | 3 | 3.1 |  |  |
| Child health insurance | |  |  |  |  |  |  | 0.288 |  |  |  |  |  |  |  | 0.553 |
|  | Yes | 2226 | 21.7 |  | 1110 | 10.8 |  |  |  | 1033 | 9.9 |  | 158 | 1.4 |  |  |
|  | No | 373 | 21.2 |  | 226 | 12.6 |  |  |  | 164 | 9.9 |  | 22 | 1 |  |  |
| ***Mothers variables*** | |  |  |  |  |  |  |  |  |  |  |  |  |  |  |  |
| Mother’s age* | | 25.3* | 5.4† |  | 24.0* | 5.0† |  | <0.001 |  | 24.2* | 5.0† |  | 24.2* | 5.1† |  | <0.001 |
|  | 11 a 24 | 1677 | 22.9 |  | 951 | 12.9 |  |  |  | 902 | 12.1 |  | 128 | 1.6 |  |  |
|  | 25 a 34 | 818 | 20.2 |  | 344 | 8.9 |  |  |  | 264 | 7 |  | 47 | 1 |  |  |
|  | 35 a 45 | 104 | 16.9 |  | 41 | 5.6 |  |  |  | 31 | 5.3 |  | 5 | 0.8 |  |  |
| Mother’s pregnancy | |  |  |  |  |  |  | 0.093 |  |  |  |  |  |  |  | 0.36 |
|  | Yes | 131 | 20.4 |  | 45 | 7.8 |  |  |  | 58 | 11.8 |  | 9 | 1 |  |  |
|  | No | 2468 | 21.7 |  | 1291 | 11.2 |  |  |  | 1139 | 9.8 |  | 171 | 1.4 |  |  |
| Number of pregnancies | |  |  |  |  |  |  |  |  |  |  |  |  |  |  |  |
|  | 1 | 1938 | 20.7 |  | 917 | 9.8 |  |  |  | 809 | 8.4 |  | 112 | 1.1 |  |  |
|  | 2 | 616 | 24.7 |  | 389 | 15.2 |  |  |  | 354 | 14.7 |  | 61 | 2.1 |  |  |
|  | 3 or more | 45 | 25.8 |  | 30 | 17.8 |  |  |  | 34 | 20.2 |  | 7 | 4.2 |  |  |
| Mother’s BMI (kg/m2) | | 25.8* | 4.3† |  | 25.3* | 4.2† |  | <0.001 |  | 24.9* | 3.7† |  | 24.6* | 3.8† |  | <0.001 |
|  | <25 | 1245 | 23.3 |  | 716 | 13.5 |  |  |  | 665 | 12 |  | 106 | 1.6 |  |  |
|  | 25 a 30 | 968 | 20.7 |  | 426 | 8.7 |  |  |  | 420 | 8.7 |  | 60 | 1.2 |  |  |
|  | >30 | 386 | 19.2 |  | 194 | 9.9 |  |  |  | 112 | 7 |  | 14 | 0.8 |  |  |
| Mother’s anemia | |  |  |  |  |  |  | <0.001 |  |  |  |  |  |  |  | 0.682 |
|  | Mild | 587 | 25 |  | 405 | 17.6 |  |  |  | 247 | 10.8 |  | 36 | 1.3 |  |  |
|  | Moderate/severe | 68 | 27.3 |  | 51 | 21.3 |  |  |  | 26 | 8.9 |  | 5 | 2 |  |  |
| Mother’s insurance | |  |  |  |  |  |  | 0.173 |  |  |  |  |  |  |  | 0.018 |
|  | Yes | 2209 | 21.2 |  | 1141 | 11.4 |  |  |  | 1059 | 10.3 |  | 163 | 1.5 |  |  |
|  | No | 390 | 21.7 |  | 195 | 9.4 |  |  |  | 138 | 7.8 |  | 17 | 0.7 |  |  |
| Marital status | |  |  |  |  |  |  | 0.184 |  |  |  |  |  |  |  | 0.032 |
|  | Single | 221 | 22.8 |  | 117 | 9.7 |  |  |  | 121 | 10.4 |  | 21 | 1.5 |  |  |
|  | Never married/cohabiting | 2111 | 21.5 |  | 1112 | 11.5 |  |  |  | 980 | 10.2 |  | 145 | 1.4 |  |  |
|  | Married/cohabiting | 267 | 21.3 |  | 107 | 8.6 |  |  |  | 96 | 7 |  | 14 | 0.9 |  |  |
| Mother’s ethnicity | |  |  |  |  |  |  | <0.001 |  |  |  |  |  |  |  | <0.001 |
|  | Spanish | 2356 | 21.2 |  | 1172 | 10.5 |  |  |  | 1009 | 9.3 |  | 129 | 1.1 |  |  |
|  | Quechua, aymara or other native languages | 243 | 28 |  | 164 | 20 |  |  |  | 188 | 20.5 |  | 51 | 5.1 |  |  |
| Mother’s education level | |  |  |  |  |  |  | <0.001 |  |  |  |  |  |  |  | <0.001 |
|  | No education/initial/primary | 416 | 25.7 |  | 216 | 12.4 |  |  |  | 368 | 25 |  | 78 | 4 |  |  |
|  | Secondary | 1357 | 23.4 |  | 769 | 13.3 |  |  |  | 612 | 9.8 |  | 82 | 1.2 |  |  |
|  | Superior technical/university | 826 | 17.8 |  | 351 | 7.6 |  |  |  | 217 | 4.6 |  | 20 | 0.6 |  |  |
| Mother’s employment status | |  |  |  |  |  |  | <0.001 |  |  |  |  |  |  |  | <0.001 |
|  | Unemployed | 1432 | 22.9 |  | 796 | 13.2 |  |  |  | 723 | 11.6 |  | 113 | 1.6 |  |  |
|  | Employed | 1167 | 20.2 |  | 540 | 8.7 |  |  |  | 474 | 8.1 |  | 67 | 1 |  |  |
| ***Home related*** | |  |  |  |  |  |  |  |  |  |  |  |  |  |  |  |
| Natural region | |  |  |  |  |  |  | <0.001 |  |  |  |  |  |  |  | <0.001 |
|  | Metropolitan Lima | 934 | 18.4 |  | 389 | 7.7 |  |  |  | 258 | 5.4 |  | 30 | 0.8 |  |  |
|  | Highland | 1004 | 26 |  | 639 | 16.5 |  |  |  | 619 | 16.8 |  | 167 | 2.3 |  |  |
|  | Jungle | 661 | 24.8 |  | 308 | 12.6 |  |  |  | 320 | 12.9 |  | 43 | 1.7 |  |  |
| Area of origin | |  |  |  |  |  |  | <0.001 |  |  |  |  |  |  |  | <0.001 |
|  | Urban | 1781 | 20 |  | 843 | 9.5 |  |  |  | 616 | 6.8 |  | 71 | 0.7 |  |  |
|  | Rural | 818 | 26.4 |  | 493 | 15.7 |  |  |  | 581 | 19.5 |  | 109 | 3.2 |  |  |
| Economic level | |  |  |  |  |  |  | <0.001 |  |  |  |  |  |  |  | <0.001 |
|  | Very low | 689 | 27.1 |  | 421 | 15.6 |  |  |  | 560 | 23 |  | 107 | 4 |  |  |
|  | Low | 821 | 23.2 |  | 477 | 15.3 |  |  |  | 347 | 10.6 |  | 46 | 1.4 |  |  |
|  | Medium | 569 | 22.4 |  | 261 | 10.5 |  |  |  | 183 | 7.6 |  | 20 | 0.6 |  |  |
|  | High | 326 | 18.4 |  | 121 | 6.4 |  |  |  | 72 | 3 |  | 5 | 0.1 |  |  |
|  | Very high | 194 | 14.3 |  | 56 | 4.1 |  |  |  | 35 | 3.7 |  | 2 | 0.5 |  |  |
| Beneficiary household of the Juntos program | |  |  |  |  |  |  | 0.062 |  |  |  |  |  |  |  | <0.001 |
|  | Yes | 266 | 23.1 |  | 147 | 13.2 |  |  |  | 239 | 21.6 |  | 55 | 4.1 |  |  |
|  | No | 2333 | 21.5 |  | 1189 | 10.8 |  |  |  | 958 | 8.8 |  | 125 | 1.1 |  |  |
| Survey year | |  |  |  |  |  |  | 0.45 |  |  |  |  |  |  |  | 0.141 |
|  | 2014 | 294 | 22.6 |  | 176 | 11.9 |  |  |  | 161 | 11.2 |  | 19 | 1.1 |  |  |
|  | 2015 | 781 | 20.9 |  | 399 | 10.8 |  |  |  | 388 | 9.8 |  | 65 | 1.7 |  |  |
|  | 2016 | 704 | 20.3 |  | 343 | 10.6 |  |  |  | 326 | 10.3 |  | 44 | 1.2 |  |  |
|  | 2017 | 820 | 22.5 |  | 418 | 10.9 |  |  |  | 322 | 8.5 |  | 52 | 1.4 |  |  |
| **n:** absolute frequency, **%:** percentage, **BMI:** Body mass index | | | | | | | | | |  |  |  |  |  |  |  |
| * mean, † standard deviation | | | | | |  |  |  |  |  |  |  |  |  |  |  |
