## Supplementary material for "Association between maternal depression and the nutritional status of children under five-years-old in Peru: An analysis of the Demographic and Health Survey 2014-2017": Table 3, and will be used for the file on the preprint site

| **Table 3. Adjusted and unadjusted association between maternal depressive symptoms and anemia in children under five years-old (Demographic and Health Survey 2014-2017)** | | | | | | | | | | | | |
| --- | --- | --- | --- | --- | --- | --- | --- | --- | --- | --- | --- | --- |
|  |  | **Bivariado** | | | | |  | **Multivariado*** | | | | |
|  |  | **Mild**† | |  | **Moderate/Severe**† | |  | **Mild**† | |  | **Moderate/Severe**† | |
|  |  | **ORm (95% CI)** | **p** |  | **ORm (95% CI)** | **P** |  | **ORm (95% CI)** | **p** |  | **ORm (95% CI)** | **P** |
| **Maternal depressive symptoms** | |  |  |  |  |  |  |  |  |  |  |  |
|  | No | 1.00 (Reference) | |  | 1.00 (Reference) | |  | 1.00 (Reference) | |  | 1.00 (Reference) | |
|  | Yes | 0.82 (0.59, 1.16) | 0.264 |  | 1.36 (0.96, 1.93) | 0.086 |  | 0.87 (0.61, 1.24) | 0.445 |  | 1.53 (0.98, 2.38) | 0.058 |
| **ORm:** odds ratio multinomial, **CI:** confidence interval | | | | | | | | | | | | |
| *Multinomial logistic regression model controlled by age of the child, sex, low birth weight, anemia of the child, chronic malnutrition of the child, child disability, health insurance of the child, age of the mother, pregnancy of the mother, number of pregnancies , Mother's BMI, mother's anemia, mother's insurance, marital status, mother's ethnicity, mother's education level, mother's employment status, natural region, area of origin, economic level, beneficiary household of the program together and survey year.. | | | | | | | | | | | | |
| †Reference category of the outcome variable for multinomial logistic regression model: no anemia. | | | | | | | | | | | | |
