## Supplementary material for "Association between maternal depression and the nutritional status of children under five-years-old in Peru: An analysis of the Demographic and Health Survey 2014-2017": Table 4, and will be used for the file on the preprint site

†Both authors contributed in the same way

**Summary**

**Objective:** To evaluate the association between maternal depression and nutritional status of children under 5 years old in Peru.

**Design:** Cross-sectional study of the Demographic and Health Survey 2014-2017. Outcomes were mild (height Z-score for age <-2 SD) and moderate/severe (<-3DE) childhood chronic undernutrition and also mild (hemoglobin <11 g/dL) and moderate/severe (hemoglobin <10 g/dL) childhood anemia. Maternal depression was assessed by the questionnaire PHQ-9. Odds ratios and their confidence intervals (CIs) were estimated by multinomial logistic regression models, considering the complex sample design.

**Setting:** Peru

**Participants:** Peruvian women of childbearing age from 15 to 49 years who live with children from 6 to 59 months.

**Results:** Maternal depression was significantly associated with a higher odds of moderate/severe chronic undernutrition in children aged 6-59 months (OR = 2.67; 95% CI 1.16-6.16).

**Conclusions:** There was evidence that maternal depression was associated with an increased risk of moderate/severe chronic undernutrition

**Keywords:** Mothers, depression, mental health, child undernutrition disorders, Peru.

There [is](https://dictionary.cambridge.org/es/diccionario/ingles-espanol/is) [no](https://dictionary.cambridge.org/es/diccionario/ingles-espanol/no) precedent [of](https://dictionary.cambridge.org/es/diccionario/ingles-espanol/of) studies that had evaluated directly the association [between](https://dictionary.cambridge.org/es/diccionario/ingles-espanol/between) [maternal](https://dictionary.cambridge.org/es/diccionario/ingles-espanol/maternal) depression [and](https://dictionary.cambridge.org/es/diccionario/ingles-espanol/and) anemia; therefore, w[e](https://dictionary.cambridge.org/es/diccionario/ingles-espanol/we) [will](https://dictionary.cambridge.org/es/diccionario/ingles-espanol/will) discuss studies [that](https://dictionary.cambridge.org/es/diccionario/ingles-espanol/that) [determined](https://dictionary.cambridge.org/es/diccionario/ingles-espanol/determined) the association between anemia and inadequate mother-child interaction. For example, Corapci et al. conducted a 5-year longitudinal study in Costa Rica, which included babies between 12 and 23 months of age and their respective mothers, that suggested a significant association between a lower quality of mother-child interaction with chronic iron deficiency (OR: 0.43, 95% CI: 0.22 to 0.87, p <0.05)^(42)^. These results differ from the marginally significant association that we found, which may be because some maternal socio demographics characteristics are different such as the maternal economic and education level. In the study mentioned mothers have a very low-low economic level and only reached incomplete high school education^(42)^. However, in our study, the maternal economic and education level are mainly low-medium and high-school or higher education, respectively.

4.	Leiva Plaza B, Inzunza Brito N, Pérez Torrejón H, et al. Algunas consideraciones sobre el impacto de la desnutrición en el desarrollo cerebral, inteligencia y rendimiento escolar. *Arch. Latinoam. Nutr.* **51**, 19.

5.	Instituto Nacional de Estadística e Informática Encuesta Demográfica y de Salud Familiar 2017. https://www.inei.gob.pe/media/MenuRecursivo/publicaciones_digitales/Est/Lib1525/index.html.

6.	Instituto Nacional de Estadística e Informática Encuesta Demográfica y de Salud Familiar 2018. https://www.inei.gob.pe/media/MenuRecursivo/publicaciones_digitales/Est/Lib1656/index1.html.

7.	Organización Panamericana de la Salud (2013) *Impacto económico de la anemia en el Perú*. *GRADE Acción Contra El Hambre*.

19. Ministerio de Salud (2018) *Plan nacional de fortalecimiento de servicios de salud mental comunitaria*. .

20. Ministerio de Salud (2016) *Salud mental comunitaria: nuevo modelo de atención*. .

21. Gelaye B, Rondon M, Araya R, et al. (2016) Epidemiology of maternal depression, risk factors, and child outcomes in low-income and middle-income countries. *Lancet Psychiatry* **3**, 973–982. NIH Public Access.

22. Husain N, Cruickshank J, Tomenson B, et al. (2002) Maternal depression and infant growth and development in British Pakistani women: a cohort study. *BMJ Open* **2**. British Medical Journal Publishing Group.

23. Harpham T, Huttly S, De Silva M, et al. (2005) Maternal mental health and child nutritional status in four developing countries. *J. Epidemiol. Community Health* **59**, 1060–4. BMJ Publishing Group Ltd.

[24. Instituto Nacional de Estadística e Informática Encuesta Demográfica y de Salud Familiar. https://proyectos.inei.gob.pe/endes/documentos.asp.](24.%20Instituto%20Nacional%20de%20Estadística%20e%20Informática%20Encuesta%20Demográfica%20y%20de%20Salud%20Familiar.%20https://proyectos.inei.gob.pe/endes/documentos.asp.)

25. Calderón M, Gálvez J & Cueva G (2012) Validación de la versión peruana del PHQ-9 para el diagnóstico de depresión. *29* **4**, 578–9.

26. Gilbody S, Richards D & Brealey S (2007) Screening for depression in medical settings with the Patient Health Questionnaire (PHQ): a diagnostic meta-analysis. *J Gen Intern Med* **29**, 578–9.

27. Sanchis F, Cortell J & Pareja H (2013) Hemoglobin point-of-care testing: the HemoCue system. *J Lab Autom* **18**, 198–205.

28. Instituto Nacional de Estadística e Informática (2018) *Informe Perú: Indicadores de Resultados de los Programas Presupuestales, 2013-2018 – Primer Semestre*.

29. World Health Organization (2015) *Levels and trends in child malnutrition*.

30. Instituto Nacional de Estadística e Informática (2018) *Lactancia y nutrición de niñas, niños y mujeres*.

[31. World Health Organization Prevalence of anaemia in children under 5 years. https://www.who.int/data/gho/data/indicators/indicator-details/GHO/prevalence-of-anaemia-in-children-under-5-years-(-).](31.%20World%20Health%20Organization%20Prevalence%20of%20anaemia%20in%20children%20under%205%20years.%20https://www.who.int/data/gho/data/indicators/indicator-details/GHO/prevalence-of-anaemia-in-children-under-5-years-(-).)

32. Instituto Nacional de Estadística e Informática (2018) *Perú: Línea de base de los principales indicadores disponibles de los objetivos de desarrollo sostenible*.

42. Corapci F, Radan AE & Lozoff B (2007) Iron Deficiency in Infancy and Mother-Child Interaction at 5 Years. 15.

43. Pullum T & Staveteig S (2017) *An Assessment of the Quality and Consistency of Age and Date Reporting in DHS Surveys, 2000-2015*. United States Agency for International Development.

[44. Instituto Nacional de Estadística e Informática Encuesta Demográfica y de Salud Familiar 2014. https://www.inei.gob.pe/media/MenuRecursivo/publicaciones_digitales/Est/Lib1211/index.html.](44.%20Instituto%20Nacional%20de%20Estadística%20e%20Informática%20Encuesta%20Demográfica%20y%20de%20Salud%20Familiar%202014.%20https://www.inei.gob.pe/media/MenuRecursivo/publicaciones_digitales/Est/Lib1211/index.html.)

[45. Instituto Nacional de Estadística e Informática Encuesta Demográfica y de Salud Familiar 2015. https://www.inei.gob.pe/media/MenuRecursivo/publicaciones_digitales/Est/Lib1356/.](45.%20Instituto%20Nacional%20de%20Estadística%20e%20Informática%20Encuesta%20Demográfica%20y%20de%20Salud%20Familiar%202015.%20https://www.inei.gob.pe/media/MenuRecursivo/publicaciones_digitales/Est/Lib1356/.)

[46. Instituto Nacional de Estadística e Informática Encuesta Demográfica y de Salud Familiar 2016. https://www.inei.gob.pe/media/MenuRecursivo/publicaciones_digitales/Est/Lib1433/index.html.](46.%20Instituto%20Nacional%20de%20Estadística%20e%20Informática%20Encuesta%20Demográfica%20y%20de%20Salud%20Familiar%202016.%20https://www.inei.gob.pe/media/MenuRecursivo/publicaciones_digitales/Est/Lib1433/index.html.)

47. World Health Organization (2006) *WHO Child Growth Standards: Length/height-for-age, weight-for-age, weight-for-length, weight-for-height and body mass index-for-age: Methods and development*.

48. World Health Organization (2019) *WHO Anthro Survey Analyser*.

| **Table 4. Adjusted and unadjusted association between maternal depressive symptoms and chronic undernutrition in children under five years-old (Demographic and Health Survey 2014-2017)** | | | | | | | | | | | | |
| --- | --- | --- | --- | --- | --- | --- | --- | --- | --- | --- | --- | --- |
|  |  | **Bivariate** | | | | |  | **Multivariate*** | | | | |
|  |  | **Mild**** | |  | **Moderate/Severe**** | |  | **Mild**** | |  | **Moderate/Severe**** | |
|  |  | **ORm (95% CI)** | **p** |  | **ORm (95% CI)** | **p** |  | **ORm (95% CI)** | **p** |  | **ORm (95% CI)** | **p** |
| **Maternal depressive symptoms** | |  |  |  |  |  |  |  |  |  |  |  |
|  | No | 1.00 (Reference) |  |  | 1.00 (Reference) | |  | 1.00 (Reference) | |  | 1.00 (Reference) | |
|  | Yes | 0.76 (0.50, 1.14) | 0.177 |  | 2.19 (0.91, 5.31) | 0.080 |  | 0.81 (0.51, 1.28) | 0.372 |  | 2.67 (1.16, 6.16) | 0.021 |
| **ORm:** odds ratio multinomial, **CI:** confidence interval | | | | | | | | | | | | |
| * Multinomial logistic regression model controlled by age of the child, sex, low birth weight, anemia of the child, chronic malnutrition of the child, child disability, health insurance of the child, age of the mother, pregnancy of the mother, number of pregnancies , Mother's BMI, mother's anemia, mother's insurance, marital status, mother's ethnicity, mother's education level, mother's employment status, natural region, area of origin, economic level, beneficiary household of the program together and survey year. | | | | | | | | | | | | |
| † Reference category of the outcome variable for multinomial logistic regression model: no chronic undernutrition. | | | | | | | | | | | | |
